# Vaping preferences of individuals who vaporise dry herb cannabis, cannabis liquids and cannabis concentrates

**DOI:** 10.1101/2022.04.23.22274223

**Authors:** Jody Morgan, Grace Gschwend, Matilda Houston, Alison Jones, Celine Kelso

## Abstract

In 2019 an estimated 200 million people aged 15-64 used cannabis, making cannabis the most prevalent illicit substance worldwide. The last decade has seen a significant expansion in the cannabis vaporiser market, introducing cannabis vaporisation as a common administration method alongside smoking and ingestion. Despite reports of increased prevalence of cannabis vaporisation there has been little research into the use of these devices. To remedy the current dearth of data in this area this study utilised an anonymous online survey of individuals who self-reported past cannabis vaporisation. The respondents (N=557) were predominantly young (<35 years) and male. Most (91.4%) stated they had ever vaped dry herb cannabis, 59.1% reported vaporisation of cannabis oil or liquids, and 34.0% reported vaporisation of cannabis concentrates. This study identifies the types of vaporisation devices (including brands and models) employed by cannabis vapers, as well as the vaporisation temperatures and puff durations commonly used for dry herb, cannabis liquids and cannabis concentrates. To the best of our knowledge, this is the first time the usual operating temperatures of these vaporisation devices and user specific consumption patterns such as puff duration have been reported for cannabis vaping. This information will allow for more realistic experimental conditions in research settings.

## Introduction

In 2019 an estimated 200 million people aged 15-64, or 4.0% of the global population, used cannabis, making cannabis the most prevalent illicit substance worldwide (UNODC 2021; WHO 2016). In Australia, 11.6% of individuals over 14 years of age reported past year cannabis use in 2019 (AIHW 2020). While cannabis can be consumed in several ways (e.g. smoking, ingestion), administration via vaporisation only became possible with the introduction of the Volcano™ table-top device in 1998, with the term vaping referring to the act of heating a mixture to the point of vaporisation, without combustion, producing an aerosol for inhalation (Hazekamp et al. 2006).

With the recent upsurge in the electronic-cigarette (e-cigarette) market (Jerzyński et al. 2021), portable vaporisers have become increasingly prevalent. There has also been a clear increase in the number of individuals reporting vaporisation as a method of cannabis administration, with an estimated 2.0% of the US adult population reporting past 30-day cannabis vaping in 2019, up from 1.0% in 2017, and a steep increase in the number of cannabis users who vaporise as their preferred administration method (14.9% in 2019 c.f. 9.9% in 2017) (Boakye et al. 2021). The recent increase in vaping is likely in part the result of cannabis vaporisation being generally regarded as healthier and more discrete than smoking (Etter 2015). However, despite the recent expansion of the cannabis vaporiser market and reports of increased use, there has been little research into cannabis vaporisation devices and methods.

Cannabis vaping can refer not only to the vaporisation of cannabis plant material (referred to throughout as dry herb cannabis) but may also relate to the vaporisation of cannabis concentrates, and cannabis containing liquids or oils, with devices often designed for optimised delivery of a specific type of cannabis product. Dry herb cannabis vaporisation employs direct use of the plant material and as such can be considered a direct substitute for smoking while vaporisation of cannabis liquids or oils most closely resembles the analogous use of e-cigarettes. Cannabis liquids are generally made by directly infusing the cannabis plant into a carrier fluid (propylene glycol (PG), vegetable glycerin (VG), or polyethylene glycol (PEG)) or by dissolving cannabis concentrates into these solvents. Cannabis liquids can be do-it-yourself (DIY) but are also commercially available as pre-made liquids and can be flavoured in the same way as nicotine-containing e-liquids. Undiluted cannabis concentrates with an oil-like consistency can also be vaporised directly in liquid form. Unlike liquids, concentrates (also known as extracts) are often referred to by their consistency (for example ‘shatter’, ‘crumble’, ‘budder’, or ‘oil’) or may be referred to by the method used to produce them. For example ‘rosin’ refers to a type of solvent-less extract generated through application of heat and pressure while butane hash oil (BHO) is a concentrate derived from a butane-based solvent extraction. Concentrates typically contain high levels of the psychoactive cannabinoid Δ9-tetrahydrocannabinol (THC) with concentrations of >90% reported (DEA 2021).

Despite the increase in cannabis vaporisation there is a dearth of information in the literature as most cannabis vaporisation studies fail to distinguish between these cannabis types. Our study aims to examine and distinguish between these three specific types of cannabis products (dry herb cannabis, cannabis liquids or oils, and cannabis concentrates) to significantly expand the literature on cannabis vaporisation, providing information on vaping preferences including: vaporisation temperatures; device types; and puff duration for the different cannabis types. This will allow for replication of accurate vaping conditions in a laboratory setting.

## Materials and Methods

Respondents were a convenience sample of adults (≥18 years old) who self-reported having ever vaporised cannabis or cannabis products. The study was approved by the University of Wollongong Human Research Ethics Committee (application number V2 15042021). Respondents were recruited via advertisements (non-paid) placed on social media platforms Facebook, Twitter and Reddit. The shared posts contained a link to the survey hosted on Qualtrics. Data was collected anonymously from May 17 to July 3, 2021. Only respondents who were ≥18 years of age and indicated that they had previously vaporised cannabis products were granted access to the survey.

The questionnaire comprised 52 questions in five blocks, including multiple choice and written responses. The full questionnaire is available in Supplementary Material 1. Block one collected information on the participants’ demographic, legality of cannabis in their location and history of cannabis use. Blocks two to four collected information specific to vaporisation of dry-herb, cannabis oil or liquids, and cannabis concentrates, respectively. Questions in these blocks included frequency of use, device types used, and methods of preparation and administration. Block five probed the participants’ preferences among products and methods of administration as well as reasons why they choose to vaporise cannabis. Most questions were not compulsory. Results were analysed using descriptive statistics, significant differences between subgroups (p<.05 with Bonferroni correction applied for post-hoc analysis) were proven using ANOVA, t-tests, or chi-square tests where relevant.

## Results and Discussion

### Respondent demographics

Of the 619 people who accessed the survey, 60 did not meet the eligibility criteria and two did not provide informed consent. Responses for the remaining 557 respondents were included in this analysis. Respondent demographics are summarised in **Table 1**. Age of respondents was positively skewed with 63.0% of respondents aged under 35 years and a median age range of 25-34 years. The majority of participants were male (75.2%, N=557). Twenty-four countries were represented in the data set with the majority residing in Australia (53.8%, N=556).

**Table 1:** Demographic characteristics of the survey respondents.

| Characteristic | Category | n (%) |
| --- | --- | --- |
| Age (N=557) | 18-24 | 195 (35.0) |
|  | 25-34 | 156 (28.0) |
|  | 35-44 | 127 (22.8) |
|  | 45-54 | 53 (9.5) |
|  | 55-64 | 18 (3.2) |
|  | 65+ | 8 (1.4) |
| Gender (N=557) | Female | 124 (22.3) |
|  | Male | 419 (75.2) |
|  | Non-binary | 12 (2.2) |
|  | Prefer not to say | 2 (0.4) |
| Country (N=556) | Australia | 299 (53.8) |
|  | United States | 141 (25.4) |
|  | United Kingdom | 33 (5.9) |
|  | Canada | 32 (5.8) |
|  | Sweden | 13 (2.3) |
|  | Germany | 7 (1.3) |
|  | Italy | 4 (0.7) |
|  | New Zealand | 3 (0.5) |
|  | Other | 24 (4.3) |

The overrepresentation of males and young adults among participants in the survey data is consistent with a 2016 survey of American cannabis users and a separate survey of American high school students which found males and younger cannabis users were more likely to report ever vaping cannabis (Lee et al. 2016; Morean et al. 2015). This may be related to higher prevalence of cannabis use (Coffey et al. 2002; Swift et al. 2001), and nicotine vape use (Dai and Leventhal 2019) among males. The positive skew of respondents’ age is consistent with nicotine vape use, which has been repeatedly demonstrated to be more prevalent among younger individuals in the US (Dai and Leventhal 2019; Mirbolouk et al. 2018).

### Cannabis usage

When asked if the primary reason for vaporising cannabis was medicinal, recreational, or both (medicinal and recreational), almost half of respondents answered ‘both’ (49.1%, N=542), just 13.5% reported vaporising cannabis for medicinal purposes only and 37.5% for recreational purposes only. A previous survey in the US found that approximately 17% of cannabis users (in states where medicinal cannabis is legal) use for medical reasons, comparable to the results presented here (Lin et al. 2016). This US survey, however, did not allow users to report both medicinal and recreational use together.

Filtering the results presented in this study to only Australian respondents (N=288); 18.0% reported vaping cannabis medicinally, 39.6% recreationally and 42.4% both recreationally and medicinally. A chi square test found that Australian users had a greater association with medicinal cannabis vaporisation than the other countries combined responses (χ^2^ (2, N=542)=15.99, p< .001). Within Australia, the National Drug and Alcohol Household Survey reported that only 6.8% of cannabis users (age 14+) were using for medicinal purposes in 2019 (16.3% both medicinal and non-medicinal, 77% non-medicinal) (AIHW 2021). The higher proportion of Australian respondents reporting medicinal use of cannabis in this study compared to the 2019 survey may be influenced by the exclusion of non-vaping cannabis users from the survey (see limitations section) or increases in use of medicinal cannabis since its legalisation in Australia in 2016 (Lintzeris et al. 2020).

The preferred method of cannabis administration among all respondents (N=445) was vaporisation (69.2%), including 10.3% who stated that they had only ever vaporised cannabis, followed by smoking (24.9%) and ingestion (5.8%). The preferred method by primary reason of use is shown in

**Figure 1A**. Across all self-reported reasons for cannabis use (recreational, medicinal, or medicinal and recreational), vaporisation was the preferred method. A chi square test identified a significant relationship between self-reported reason for use and preferred administration method (χ^2^ (6, N=445)=22.84, p< .001). Assessment of the contributors indicated that recreational users are associated with a higher preference for smoking and lower preference for vaping (Supplementary Material 2 Table SM1). Conversely, medicinal and recreational users are associated with a higher preference for vaping and lower preference for smoking. Medicinal only use is associated with a greater preference for ingestion. This aligns with the 2018-2019 Cannabis as Medicine Survey which reported that Australians who use cannabis for therapeutic reasons demonstrated a significant preference for oral (ingestion of edibles) or vaporised routes of administration over combustion methods (joints, water pipes and glass pipes) (Lintzeris et al. 2020).

**Figure 1:**
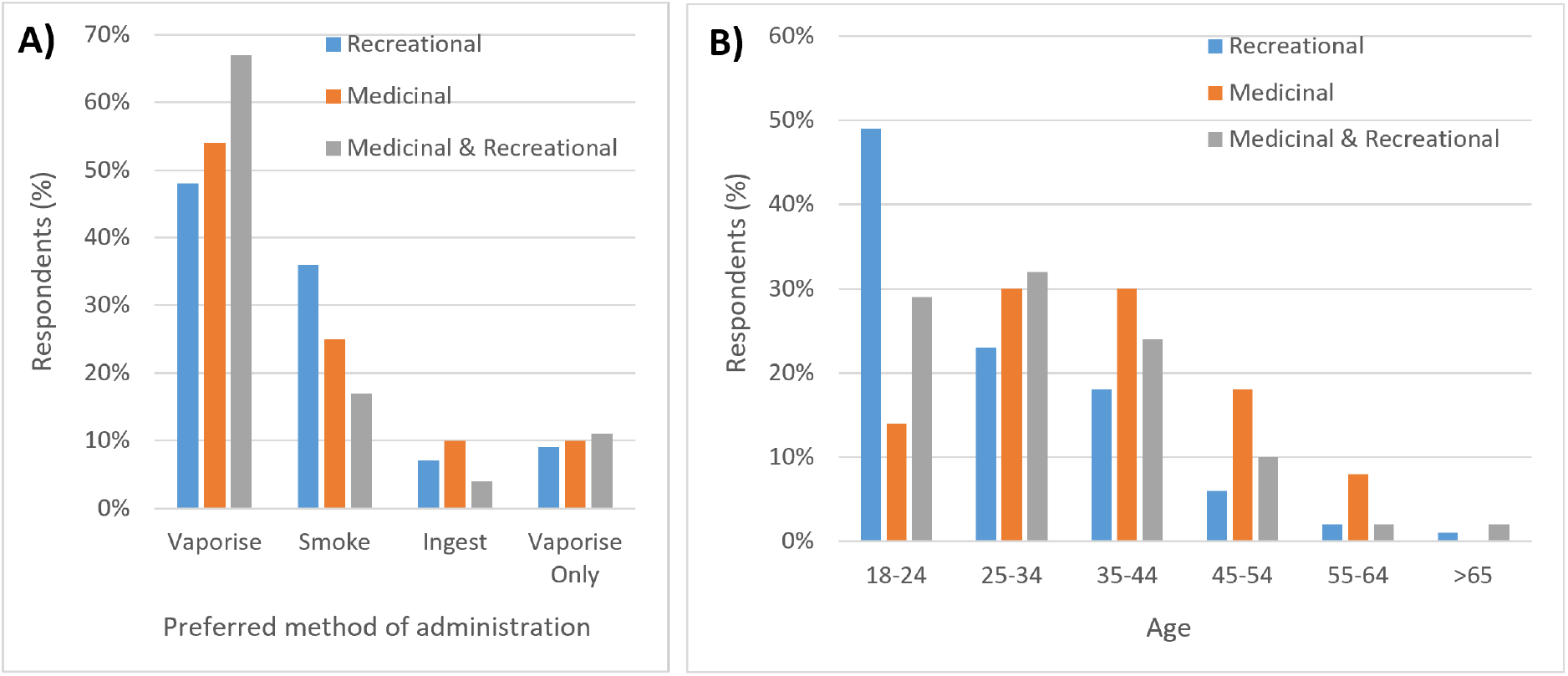
**A)** Reason of cannabis use and self-reported preferred method of administration among respondents (N=445). **B)** Reason of cannabis use and age distribution among respondents (N=542).

Several studies have previously reported medicinal cannabis users are likely to be older individuals (Lin et al. 2016; AIHW, 2021). This is supported by the results of this survey (**Figure 1B**) which found a significantly greater proportion of 18-24 year-olds in the recreational category and a greater proportion of medicinal users in older age categories, particularly the over 35 year-old age groups (χ^2^ (10, N=542)=48.55, p< .001).

While the length of time that participants had been using cannabis by any method other that vaporisation appeared to be randomly distributed, the period they had been vaporising cannabis was skewed towards shorter timeframes (**Figure 2A**). More than 85% of respondents (N=550) reported vaping cannabis for five years or less, with the majority (50.2%) self-reporting cannabis vaporisation for between one and five years. In contrast, 10.9% of respondents reported cannabis vaporisation for five to ten years, and only 3.6% for more than ten years. The increased prevalence of recent cannabis vaping is likely due to the growth in the cannabis vaporiser market in the last decade, which has made cannabis vaporisation more accessible and convenient (Wadsworth et al. 2022).

**Figure 2:**
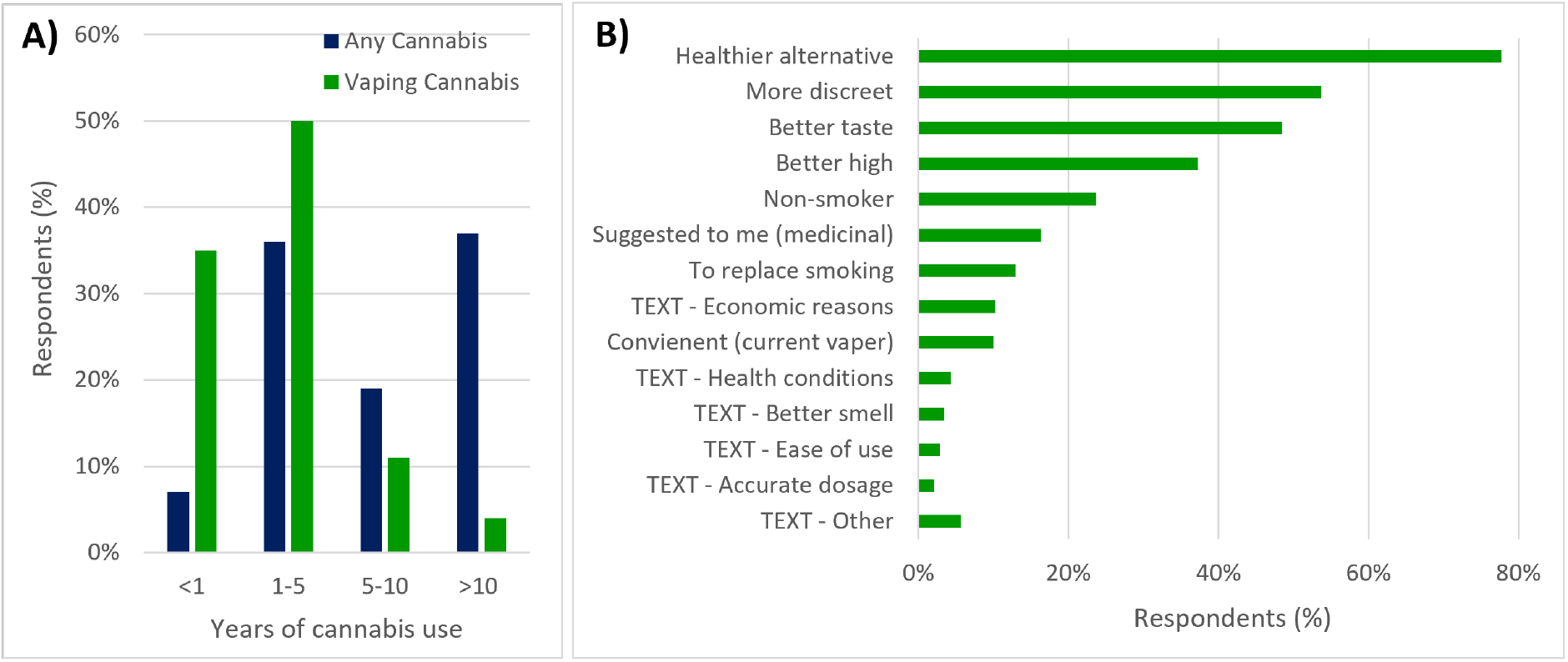
**A)** Surveyed participants (N=550) length of cannabis use as a function of intake method (cannabis vaporisation vs any other intake method). **B)** Self-reported reasons why respondents choose to vape cannabis products, respondents could select multiple options (N=440). “TEXT” identifies responses provide as a freeform text response.

Respondents gave many reasons as to why they choose to vape cannabis products, respondents were able to select multiple options provided for this question and provide an additional text response (**Figure 2B**). The most common response was that vaporising cannabis is considered healthier than smoking with more than three quarters of respondents citing this as a reason they choose to vaporise their cannabis (77.7%, N=440). More discretion and better taste were the next most common reasons cited by 53.6% and 48.4% of respondents, respectively. More than a third (37.3%) of participants reported that vaping gives a ‘better high’ than smoking. These results are consistent with a 2016 survey of US cannabis vape users (N=2910) who reported perceptions that vaporisation is healthier than smoking (72%), gives a better taste (55%), and provides a better high (∼50%) (Lee et al. 2016).

One hundred and seventeen participants provided additional text responses to this question. The most cited ‘other’ reason was that vaping is more efficient or economical than smoking, producing the same effects at lower doses or otherwise reducing cannabis consumption (n=45). Other common reasons for vaping rather than smoking included treatment of underlying health conditions (n=19), a better or less noticeable smell (n=15), convenience/ease of use (n=13), and more accurate or controlled dosage (n=9). Consistent with the results of this survey, past research identified the main advantages of vaporisation perceived by cannabis users were: perceived health benefits, better taste, no smell/more discreet, and more effect for the same amount of cannabis (Malouff et al. 2014; Morean et al. 2017).

### Cannabis type preferences

Of the 535 respondents, 91.4% stated they had ever vaped dry herb cannabis, 59.1% had vaped cannabis oil or liquids, and 34.0% had vaped cannabis concentrates. Cannabis type by age group is shown in **Figure 3A**. Fifty six percent (N=535) of respondents reported having vaped more than one type of cannabis product with 26.9% reporting they had vaporised two of the available cannabis types and 28.8% reporting they had vaporised all three types. The number of different types of cannabis vaporised by age group is shown in **Figure 3B**. Respondents who had vaped concentrates were more likely to have also vaped another type of cannabis product with 99.5% of concentrate vapers reporting also vaping another type of cannabis. In contrast 88.3% of respondents reporting vaporisation of cannabis liquids or oils and only 59.3% of respondents reporting vaporisation of dry herb cannabis also reported vaporisation of at least one other type. Respondents were also asked about their preferred cannabis type (N=422), 81.0% reported dry herb cannabis as their preferred type, 8.5% reported cannabis oil or liquid and 10.4% reported cannabis concentrates. In a previous study it was found that among those who reported ever vaporising cannabis within a medicinal cannabis group, 97.5% had used dry herb, 19.6% had vaped resin, 18.8% vaped BHO, 14.6% vaped oil, and 1.7% vaped an alcohol or carbon dioxide extract (Shiplo et al. 2016). A study by Etter et al also found that dry herb cannabis was the most commonly vaporised cannabis product with 70.9% of respondents reporting they had vaped dry herb, 27.2% had vaped cannabis ‘oil’, and 16.4% had vaped cannabis wax (N=55) (Etter 2015).

**Figure 3:**
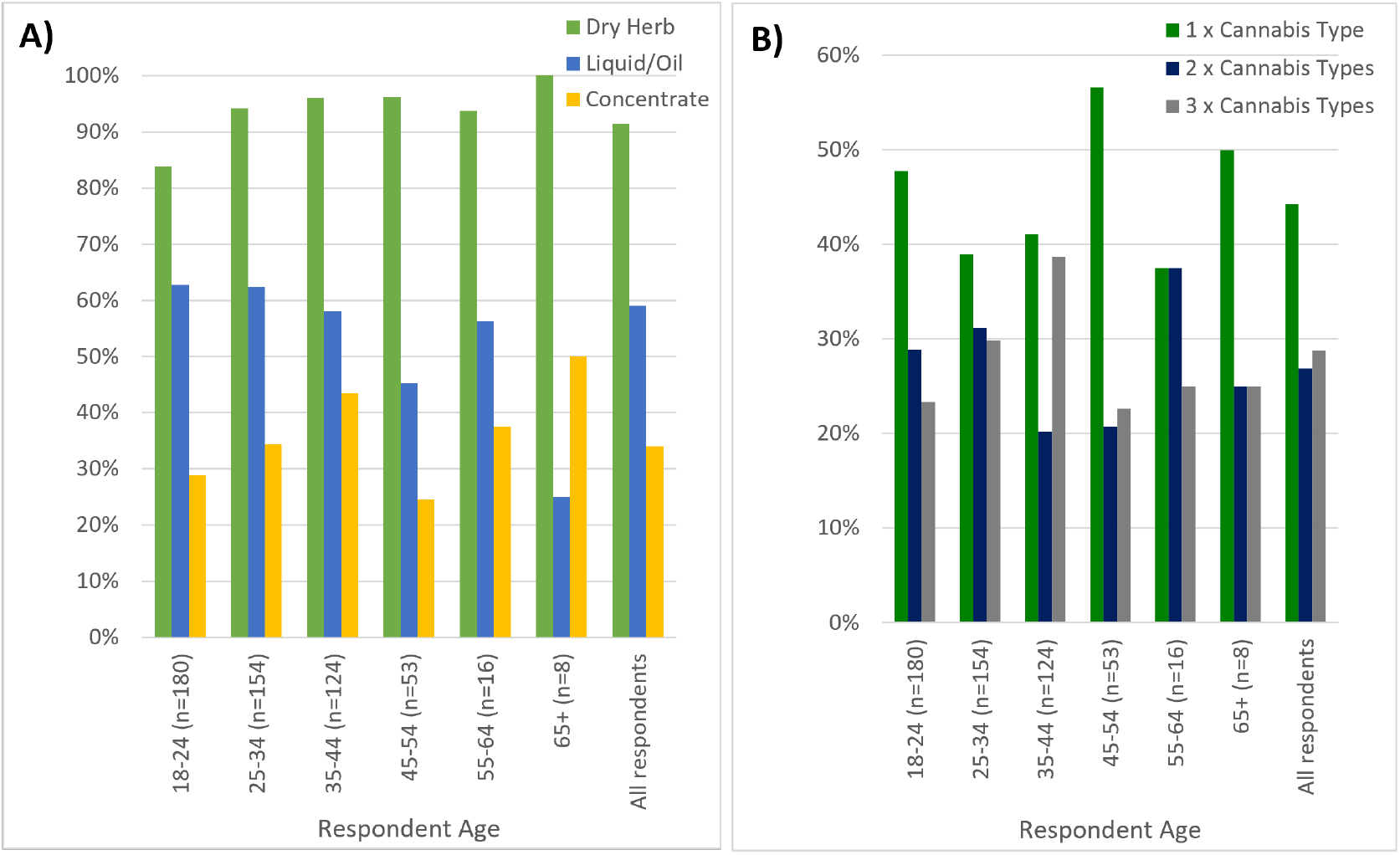
**A)** Cannabis type (dry herb vs liquid/oil vs concentrate) by age group. **B)** Number of different types of cannabis (one vs two vs three) vaporised by age group.

### Cannabis device preferences and brands

Portable vaporisers were the most common device used to vaporise all three cannabis types: dry herb (93.6%); cannabis liquids or oils (92.4%); and concentrates (60.3%) (**Figure 4**). In contrast, the use of desktop devices was much lower with reported desktop vaporiser usage rates between 15-33% across all three types, while dab-rigs were used to vaporise mostly concentrate (60.3%) and to a lesser extent e-liquids (34.1%) (dab-rig was not an available option for dry herb cannabis vaporisation).

**Figure 4:**
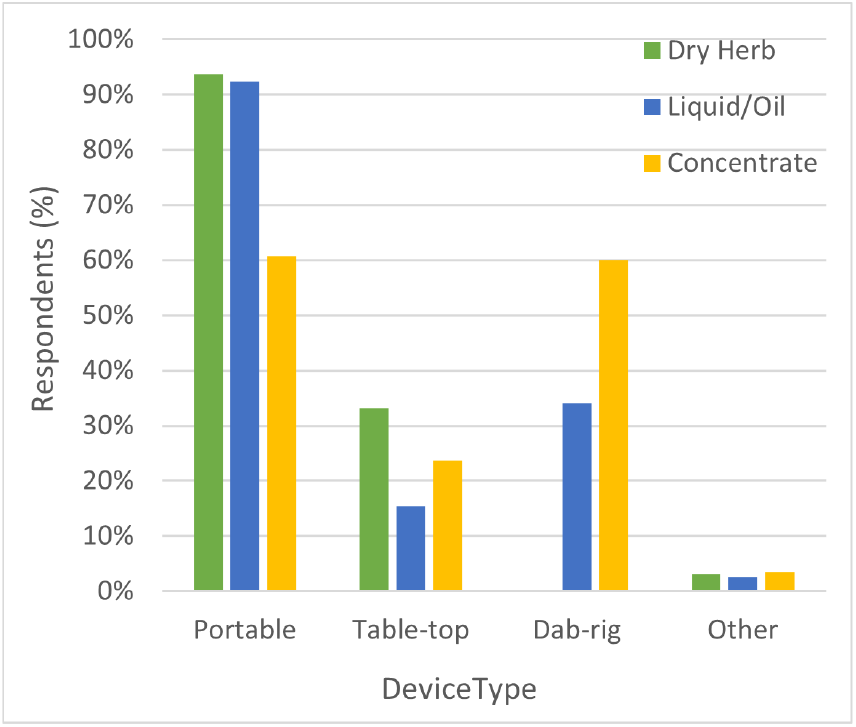
Device types reported by type of cannabis. Respondents were able to select more than one type of device. Dab-rig was not an available selection for dry herb cannabis.

Respondents who reported vaporisation of cannabis in multiple ways were asked about their preferred style of device. Seventy-nine percent of respondents (N=376) claimed to prefer vaping cannabis in a portable (handheld) vaporiser, 15.7% preferred tabletop vaporisers, and 5.3% preferred dab-rigs. A preference for portable devices to vaporise cannabis has been previously reported in the literature (Lee et al. 2016).

Chi square tests indicated no significant association between reason of cannabis use and preferred product (χ^2^ (4, N=422)=3.90, p=.41) or style of device (χ^2^ (4, N=376)=9.06, p=.060). There was, however, a strong association between preferred cannabis type and preferred device used with that product (χ^2^ (4, N=372)=142.2, p< .001). Examination of the chi square contributors (Supplementary Material 2 Table SM2) demonstrated the following associations: 1) cannabis concentrates are strongly associated with a higher preference for dab-rigs and lower preference for portable devices; 2) vaporisation of oils or liquids is associated with a lower preference for tabletop devices. These associations can be attributed to the style of the device, which are optimised for use of specific products (Chadi et al. 2020).

Survey respondents were asked to list brands of vaporisation devices they had used to vaporise cannabis for each of the three cannabis types. All text responses were analysed and identifiable brands and models were counted by cannabis type. Brands which were identified by at least five respondents are shown in **Figure 5** along with specific models, where mentioned. For brands of vaporiser used to administer dry herb cannabis, 675 identifiable text responses were collected identifying 73 different brands. The most frequently mentioned brands were Storz & Bickel (19.1%), DynaVap (17.5%), Arizer (16.0%), PAX (8.0%) and Healthy Rips (4.9%). For respondents who reported vaporisation of cannabis liquids or oils there were 188 identifiable text responses across 78 different brands. The most frequently mentioned brands for cannabis liquid or oil vaporisation were Yocan (6.9%), Vaporesso (6.4%), Storz and Bickel (5.9%), Puffco (5.3%) and Dynavap (5.3%). There were 121 identifiable text responses identifying brands used to vaporise cannabis concentrates across 48 different brands. The most frequently mentioned brands were Dynavap (14.0%), Storz and Bickel (9.9%), Puffco (6.6%), Arizer (6.6%) and Grenco (5.8%).

**Figure 5:**
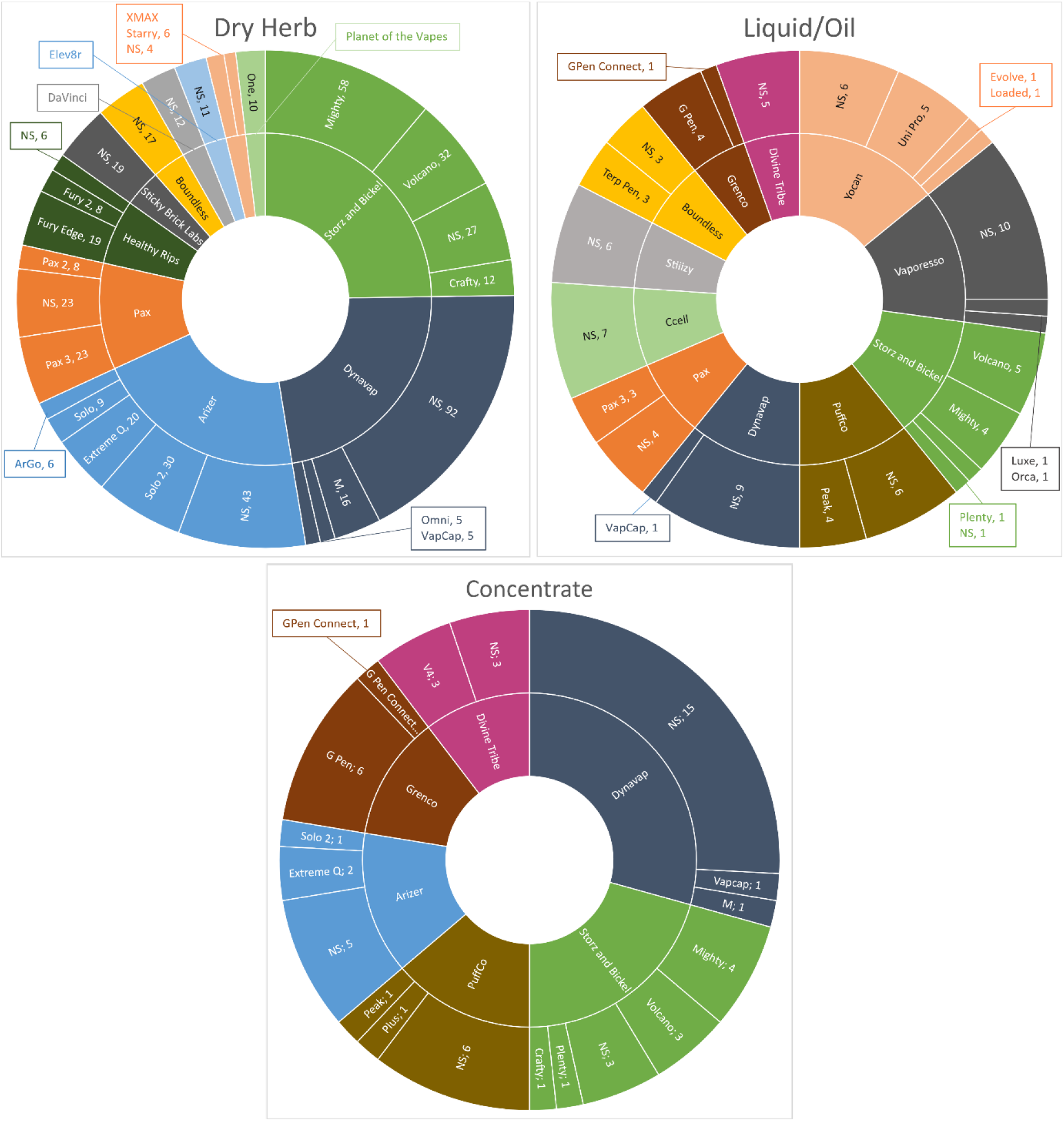
Vaporiser brands (and models if specified) identified from text responses describing devices used to vaporise different types of cannabis. Only brands with at least five responses are included in the graphic. NS = Model not specified.

To date there has only been one study examining brands used to vaporise cannabis (Etter 2015). This exploratory study found similar results among tabletop vaporisers, with the Storz & Bickel Volcano and Plenty models being the most prevalent. The same study also reported on 14 portable vaporiser brands, of which six were also observed in this survey: PAX, DaVinci, Magic Flight Launch Box, GPen, Da Buddha, and Iolite. Differences in the brands identified are likely due to the small sample size employed in the previous study and the expansion and evolution of the vaporiser market since 2014.

All text responses which provided detail on device models, or brands if only one style of device was available, were classified according to the options shown in **Figure 6**. This analysis suggests that portable vaporisers specifically designed to vaporise cannabis (‘portable cannabis vape’) were the most common classification for vaporisation of dry herb cannabis followed by thermal extraction devices, such as the Dynavap. The most reported device type for vaporising cannabis liquids and oils was vaporisers consisting of a 510 battery which takes a cannabis cartridge (usually pre-filled) followed by portable vaporisers designed for cannabis and portable vaporisers designed for e-liquids (‘portable e-cigarette’). The increased use of devices designed to vaporise nicotine-based e-liquids for cannabis liquids/oils is to be expected as these devices are likely compatible with several PG/VG based liquids containing THC or cannabidiol (CBD) commercially available. Reported device types for vaporisation of cannabis concentrates were most commonly classified as portable vaporisers designed for cannabis, dab-rigs and thermal extraction devices.

**Figure 6:**
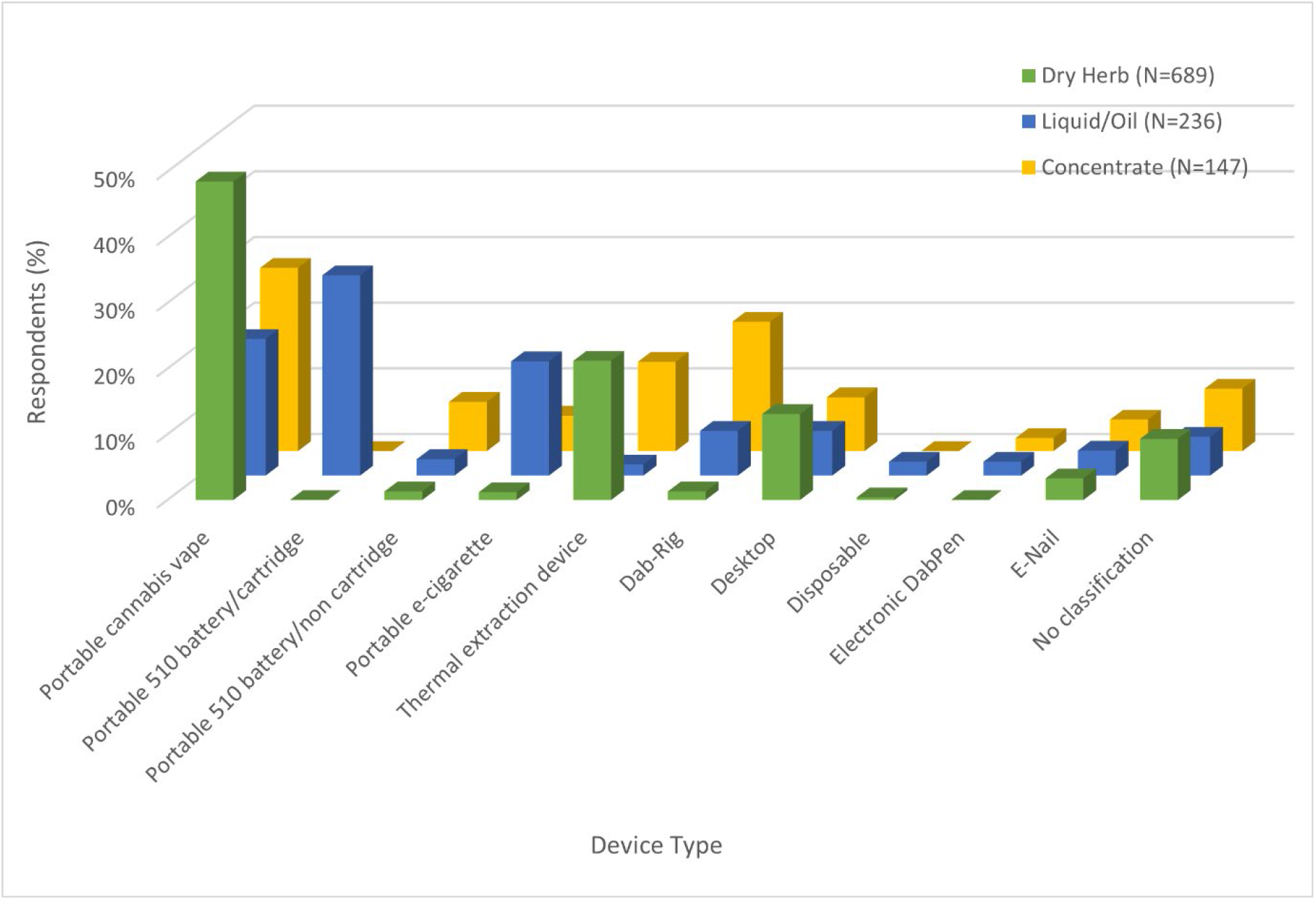
Style of device reported per type of vaporised cannabis product. Vaporisers were classified into the categories listed. Devices were classified as ‘cannabis’ or ‘e-cigarette’ devices based on either their description on the vendor website or whether they were available from websites which primarily sell cannabis or nicotine products. Brands which sell multiple styles of device were excluded from this analysis unless the model was provided in the text response.

The popularity of DynaVap and similar devices in this study was not anticipated. These ‘thermal extraction devices’ are externally heated with a flame (usually butane-sourced) rather than beingfind electric, but act as vaporisers as they heat the cannabis to the point of vaporisation without combustion. Other examples of gas or torch powered vaporisers respondents mentioned within this study included the Stickybrick, Elev8r and Vaporgenie. Use of gas-powered devices to vaporise cannabis has been speculated within one other study, however, the prevalence of use was unclear (Morean et al. 2017) with no other thermal extraction devices explicitly mentioned anywhere in the cannabis literature. DynaVap devices retail for approximately $100 AUD and have had a rapid uptake since the establishment of the brand in 2015 (Dynavap 2022). Further studies should be carried out to examine the popularity of these devices and determine if there is a trend towards butane heated devices or if the DynaVap is an anomaly in a market saturated with electronic vaporisers.

### Amount of Cannabis Vaporised

Reported amounts of cannabis vaporised per month by respondents varied widely (**Figure 7**). All responses were converted to grams before data analysis. Outliers less than three times the interquartile range (IQR) below the first quartile, or three times IQR greater than the third quartile were excluded from the data set. The average mass (outliers removed; N=366) of dry herb cannabis vaporised by respondents per month was 15.0 ± 17 g, with a median value of 9 g. The average volume (outliers removed; N=111) of cannabis liquid/oil vaporised by respondents per month was 12.9 ± 21 mL with a median value of 3 mL. The average mass of cannabis concentrates (outliers removed; N=83) vaporised by respondents per month was 4.6 ± 7 g with a median value of 1 g.

**Figure 7:**
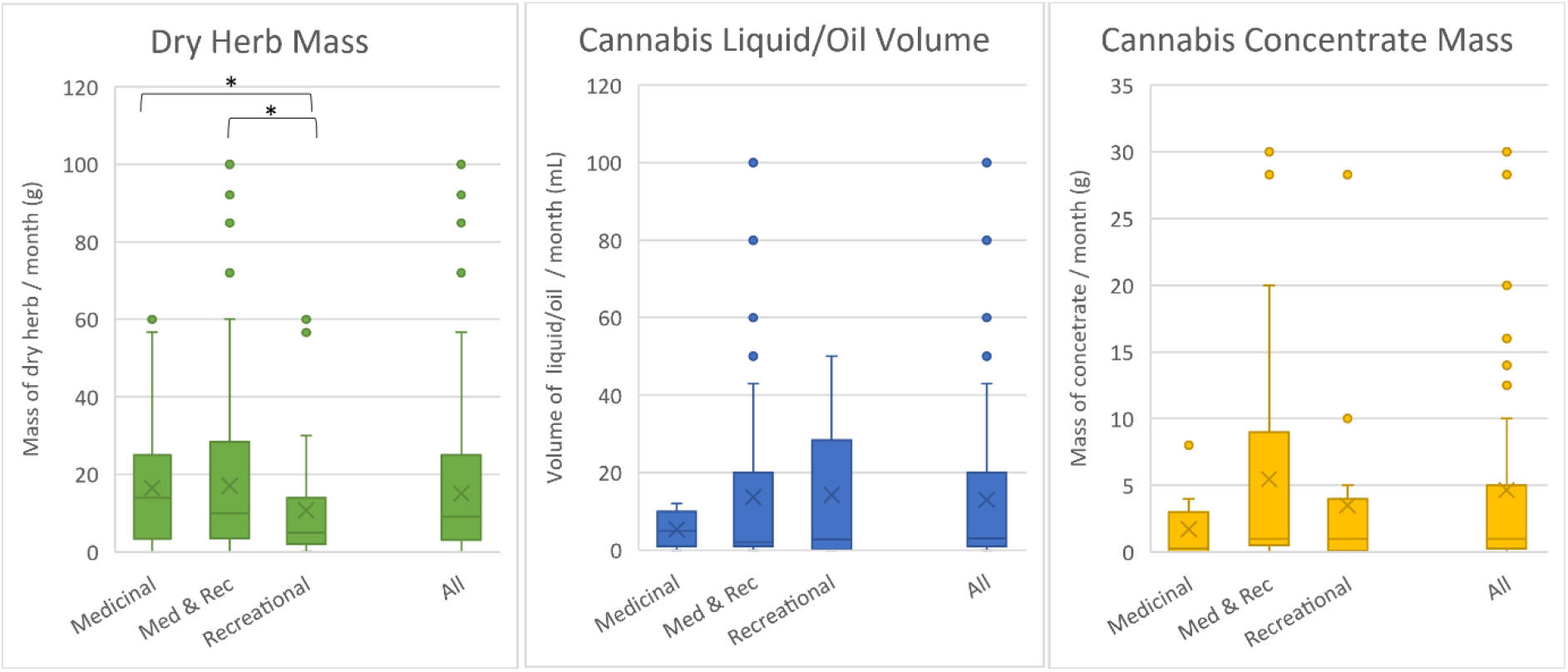
Reported amount of cannabis consumed per month. Data was analysed by segregating respondents self-identifying as medicinal only; recreational and medicinal; or recreational only users; and by providing the same data as an all respondent combined comparison. All plots were obtained from the data set after outliers were removed. * indicates p< .017 (Bonferroni correction)

These results were analysed (p< .017 (Bonferroni correction); ANOVA) by self-reported reason for cannabis vaporisation (**Figure 7**). The results for dry herb cannabis indicated that among the respondents to this survey recreational users vaped significantly less dry herb per month than medicinal users and less than medicinal and recreational users (Supplementary Material 2 Table SM3). This is likely due to the lower frequency of use among recreational users. This trend did not hold for cannabis liquid/oil or cannabis concentrates (Supplementary Material 2 Table SM3).

### Puff Duration

Respondents were asked to report their puff duration time (in seconds) for each of the different cannabis types. For all cannabis types the responses were bimodally distributed (see Supplementary Material 2 Figure SM1). It is important to note that the maximum puff duration that could be selected was ten seconds, it is possible individuals that chose ten seconds may have preferred to select a longer time. Mean puff durations of 6.3 ± 2.6 seconds (N=456), 5.1 ± 2.5 seconds (N=293) and 5.9 ± 2.6 seconds (N=163) were reported for dry herb, cannabis liquid/oil and cannabis concentrate vapers, respectively (**Figure 8A**). There was a significant difference (p< .017 (Bonferroni correction); ANOVA Supplementary Material 2 Table SM4) for the puff duration between dry herb vaporisation and liquid/oil vaporisation and between liquid/oil vaporisation and concentrates (**Figure 8A**).

**Figure 8:**
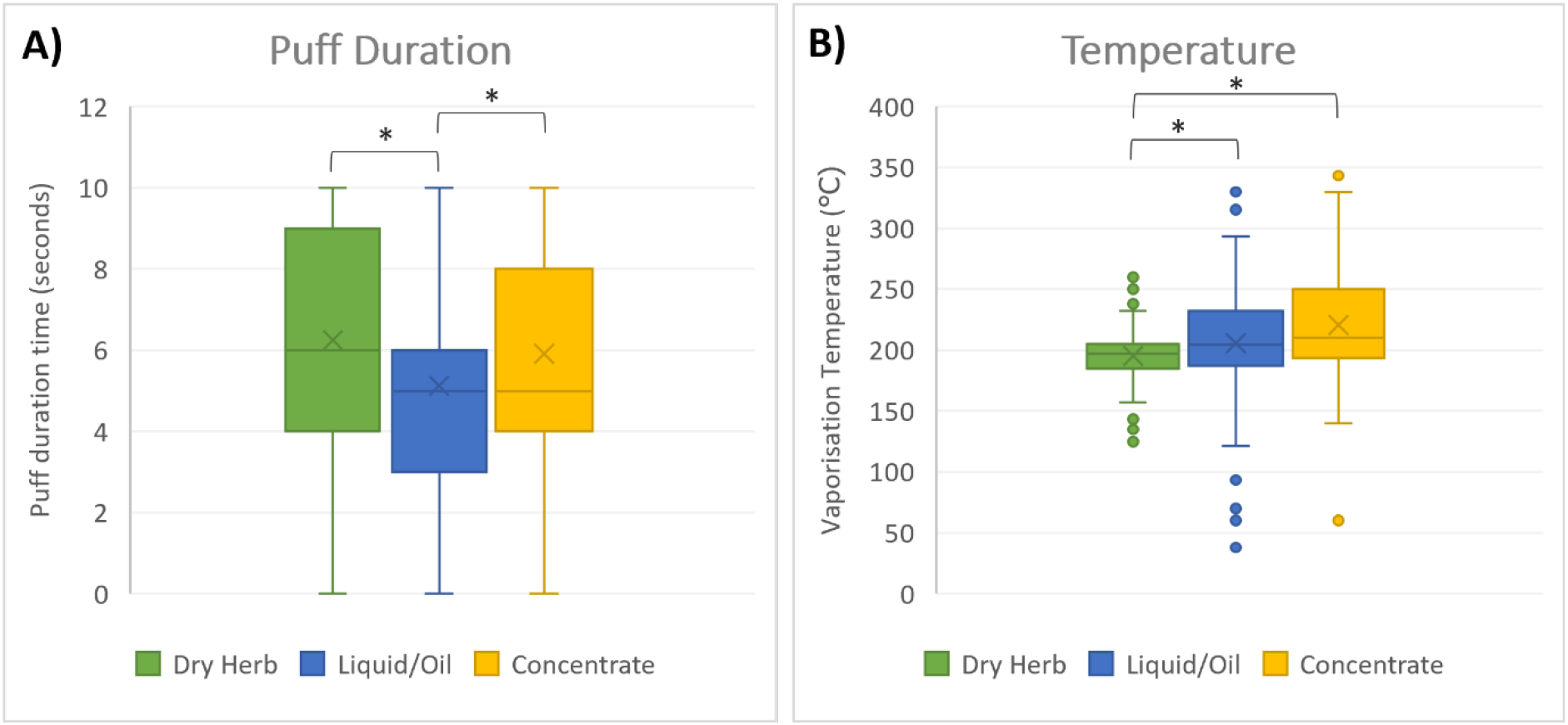
Cannabis vaper preferences: **A)** Box and whisker plot of reported puff duration (seconds) for individuals vaporising different types of cannabis. **B)** Box and whisker plot of reported temperature (°C), outliers removed, used by respondents to vaporise different types of cannabis. * indicates p< .017 (Bonferroni correction)

There is currently no available information in the scientific literature examining puff duration and inhalation patterns used for the vaporisation of cannabis. Online blogs and device instruction manuals suggest inhaling slowly, for as long as is comfortable without coughing (Monroe Blvd 2021; Weedmaps 2020). It is likely that the ten second maximum puff duration available for selection, which was selected based on the 3 second puff common in e-cigarette vaporisers and a 95^th^ percentile of 5.6 seconds (McAdam et al. 2019), was not adequate to capture the puff duration for all cannabis vapers. Puff pattern may also affect the reported puff duration, for example, when using a dab-rig the user may continually ‘hit’ the dab-rig, taking continual puffs until all the concentrate has been consumed to prevent any vapour loss. The shorter puff duration for cannabis liquids/oils suggests these are likely being inhaled in a similar pattern to e-liquids in e-cigarettes. This is likely because the cannabis liquid cools the heating coil rapidly allowing for short inhalational puff patterns with large rests between inhales and multiple heating/cooling cycles on the one sample. In contrast to dry herb and concentrate vaporisers where the entire sample in the vaporiser may be consumed in a single heating/cooling cycle. Future observational studies would be useful to fill this gap in knowledge.

### Temperature

Respondents were asked to report the temperature they used to vaporise each of the three cannabis types. Temperatures could be reported as °C or °F, all responses were converted to °C before data analysis. Outliers were removed as described above. Vaporisation temperatures for all cannabis types were normally distributed about the mean (Supplementary Material 2 Figure SM2). Respondent reported vape operating temperatures ranged from 125 to 260 °C with a mean of 195.3 ± 20 °C (N=337, outliers removed) for dry herb cannabis, from 38 to 330 °C with a mean of 205.7 ± 59 °C (N=78, outliers removed) for cannabis liquids or oils and from 60 to 343 °C with a mean of 220.6 ± 50 °C (N=71, outliers removed) for cannabis concentrates (**Figure 8B**). An ANOVA analysis (Supplementary Material 2 Table SM5) determined a significant difference (p< .017 (Bonferroni correction)) between the reported temperatures for dry herb vaporisation and cannabis liquid/oil vaporisation and between dry herb and concentrates (**Figure 8B**).

For dry herb cannabis and cannabis concentrates, a secondary analysis was performed for those who specified use of a single device type (**Figure 9**; individuals reporting use of more than one device type were excluded from this analysis). Mean vaporisation temperatures of 211.0 ± 31 °C (N=5) and 194.8 ± 20 °C (N=192) were reported for devices vaporising dry herb using only table top or only portable devices, respectively. While mean temperature of 205.9 ± 31 °C (N=21), 191.7 ± 10 °C (N=3), and 233.9 ± 64 °C (N=21) were reported for cannabis concentrates vaporised using portable, tabletop or dab-rig devices, respectively. Small variations between average temperatures across device types were observed but these were not significant (ANOVA; Supplementary Material 2 Table SM6). The higher (non-significant) temperature for dab-rigs is not unexpected. Tabletop and portable vaporisation devices commonly have predefined temperature settings available, for example, the Volcano™ tabletop vaporiser is designed to operate between 130-230 °C (Australian Vaporizers 2022), whereas glass dab-rigs are typically heated with a butane torch until hot enough to vaporise the extract. Similarly, electronic dab-rigs such as the Puffco Peak, have a higher operating range of 230-315 °C (PuffCo 2020).

**Figure 9:**
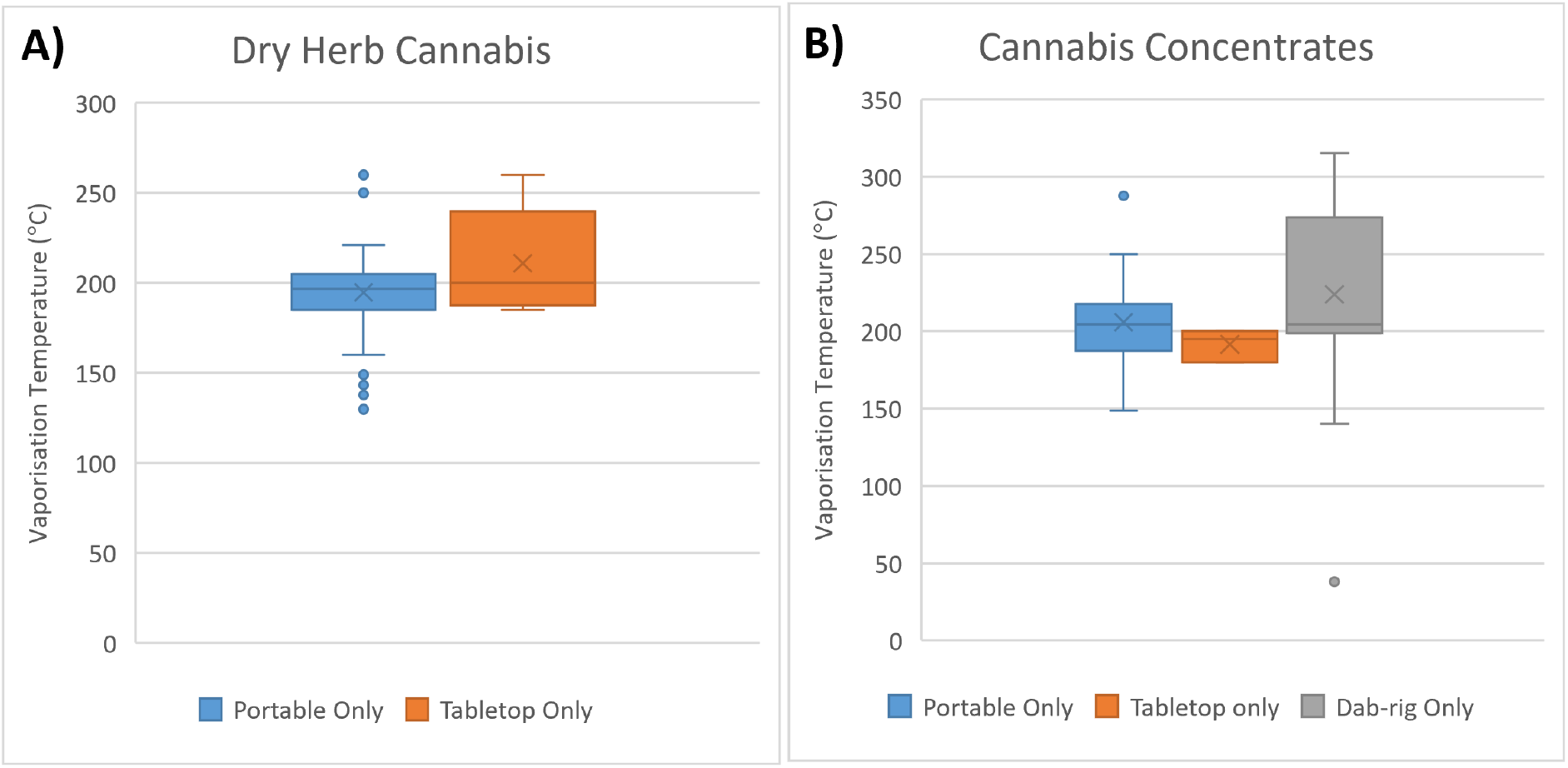
Box and whisker plot for **A)** dry herb cannabis only vaporisation temperature, by device type (tabletop (N=5); portable (N=192)) and **B)** concentrates only vaporisation temperature, by device type (portable (N=21); tabletop (N=3); dab-rig (N=21)). Individuals who reported use of more than one type of device were excluded from this analysis.

This survey is the first, to our knowledge, which has gathered information on the usual operating temperatures of vaporisation devices to administer cannabis. It is important to note that the temperatures described in this study were those reported by users. Recent research articles which employ vaporisation temperatures of up to 400 °C (Czégény et al. 2021), do not appear to reflect realistic dry herb cannabis vaporisation conditions. Knowing that cannabis vapers will not operate vaporisers at temperatures greater than 230 °C for dry herb cannabis and 300 °C for liquid/oil and cannabis concentrates will assist future research by creating more realistic vaporisation models and ensure pyrolysis products generated at extreme temperatures are not unreasonably generalised to vaporiser use.

### Limitations

The participants in this survey were a self-selected sample of the broader population of cannabis vaporiser users. The advertising method used (social media and Reddit) may have influenced the type of respondents. More specifically, posting the survey on Reddit forums habituated by vaporiser enthusiasts (subset of general population of cannabis users) may have skewed the demographic to respondents who prefer vaporisation over other methods of administration of cannabis. Previous surveys of recreational users generally show a preference for smoking (Gould et al. 2019), while even among medicinal users, the preference for vaporisation is not as high as 60% (Lintzeris et al. 2020).

The survey was posted and advertised in English which is likely responsible for the high proportion of respondents from Australia, North America, and the United Kingdom. Due to the limited number of respondents, the average response across all countries was reported here. The legality of cannabis and that of electronic vaping devices in the region where respondents reside, ease of use or acquisition, associated high, and/or onset of effects may also have influenced responses, particularly the types of cannabis products and vaporisation devices available to/preferred by respondents.

The question examining puff duration had an upper limit of ten seconds available for selection. It was not expected that users would inhale for more than ten seconds, however, the bimodal distribution of responses indicates this presumption was likely incorrect. Observational studies of cannabis vaporisation may be employed for more accurate results than is achieved by self-reporting.

It is important to note there was some confusion among a small number of respondents as to the type of cannabis the questions were referring to, despite the definition and sample images provided for all types. This was particularly relevant for cannabis liquids/oils vs cannabis concentrates, with several text-based responses in the cannabis liquids/oils section describing cannabis concentrates or extracts. Researchers need to ensure that nomenclature around cannabis types is clear with specific definitions to allow respondents to improve the quality of responses.

## Conclusions

This study has provided evidence-based usage rates and preferences for dry herb cannabis, cannabis liquids/oils and cannabis concentrates, being one of the first studies of its kind to assess these cannabis types separately. The study identified the types of vaporisation devices, brands and models used most commonly with each of the cannabis types. Additionally, the vaporisation temperatures and puff durations commonly used for dry herb, cannabis liquids and cannabis concentrates were reported for the first time. These evidence-based vaporisation settings, used by vapers in the community across the different product types, will allow future realistic vaping modelling in the laboratory to further assess the content of these products. Users’ motives for choosing certain types of cannabis products or vaporisation devices were not examined in this survey and should be investigated further in future.

## Data Availability

All data produced in the present study are available upon reasonable request to the authors.

## Notes

### Competing Interest Statement

The authors have declared no competing interest.

### Funding Statement

This study did not receive any funding.

### Author Declarations

The Human Research Ethics Committee of The University of Wollongong gave ethical approval for this work (application number V2 15042021).

