## Supplementary Material for "Vaping preferences of individuals who vaporise dry herb cannabis, cannabis liquids and cannabis concentrates"

**Running title:** Cannabis Vaper Preferences

### Abstract

In 2019 an estimated 200 million people aged 15-64 used cannabis, making cannabis the most prevalent illicit substance worldwide. The last decade has seen a significant expansion in the cannabis vaporiser market, introducing cannabis vaporisation as a common administration method alongside smoking and ingestion. Despite reports of increased prevalence of cannabis vaporisation there has been little research into the use of these devices. To remedy the current dearth of data in this area this study utilised an anonymous online survey of individuals who self-reported past cannabis vaporisation. The respondents (N=557) were predominantly young (<35 years) and male. Most (91.4%) stated they had ever vaped dry herb cannabis, 59.1% reported vaporisation of cannabis oil or liquids, and 34.0% reported vaporisation of cannabis concentrates. This study identifies the types of vaporisation devices (including brands and models) employed by cannabis vapers, as well as the vaporisation temperatures and puff durations commonly used for dry herb, cannabis liquids and cannabis concentrates. To the best of our knowledge, this is the first time the usual operating temperatures of these vaporisation devices and user specific consumption patterns such as puff duration have been reported for cannabis vaping. This information will allow for more realistic experimental conditions in research settings.

### Supplementary Material 1 - QUESTIONNAIRE

**BLOCK 1 - Eligibility Questions**

1. What is your current age?
    a. <18
    b. 18-24
    c. 25-34
    d. 35-44
    e. 45-54
    f. 55-64
    g. >65

*[If <18 respondents sent directly to the end of the survey]*

2. What is your gender?
    a. Male
    b. Female
    c. Non-binary
    d. Prefer not to disclose

3. What country are you in? [TEXT ANSWER]

4. Is cannabis use legal in the state/country you are in?
    a. No, it is illegal
    b. It is legal for medicinal use only
    c. It is legal for medicinal and recreational use
    d. Unsure

5. Have you ever used cannabis (marijuana) in any form? This includes smoking, eating or vaping of cannabis flower, hash, concentrates, oils, THC or CBD.
    a. Yes
    b. No

6. *[If yes to Q5 include this question]* How long have you been using cannabis for? This includes smoking, eating or vaping cannabis in any form.
    a. less than 12 months
    b. 12 months to 5 years
    c. 5 - 10 years
    d. more than 10 years

7. Have you ever vaporised cannabis (marijuana) in any form? This includes vaporising cannabis flower, hash, concentrates, oils, THC or CBD in an e-cigarette style device, vape-pen, tabletop vaporiser or dab-rig.
    a. Yes
    b. No

    *[If no to Q7 respondents sent directly to the end of the survey]*

8. *[If yes to Q7 include this question]* How long have you been vaping cannabis for? This question is specifically referring to vaporising cannabis and excludes eating and smoking.
    a. less than 12 months
    b. 12 months to 5 years
    c. 5 - 10 years
    d. more than 10 years

**BLOCK 2 - Cannabis Usage General Questions**

9. When you smoke cannabis you do this primarily:
    a. For medicinal purposes
    b. For recreational purposes
    c. For both medicinal and recreational purposes
    d. I have never smoked cannabis

10. When you ingest (eat/drink) cannabis you do this primarily:
    a. For medicinal purposes
    b. For recreational purposes
    c. For both medicinal and recreational purposes
    d. I have never ingested cannabis

11. When you vaporise cannabis in any form (this includes vaporising cannabis flower, hash, concentrates, oils, THC, CBD or flavoured cannabis-containing e-liquids) in an e-cigarette style device, vape-pen, tabletop vaporiser or dab-rig you do this primarily:
    a. For medicinal purposes
    b. For recreational purposes
    c. For both medicinal and recreational purposes

**BLOCK 3 - Dry Herb Cannabis**

12. Have you ever vaporised dry herb cannabis (flower/plant material) in any device? This question is specifically referring to vaporising cannabis plant material only and excludes vaporisation of cannabis concentrates, oils or liquids.

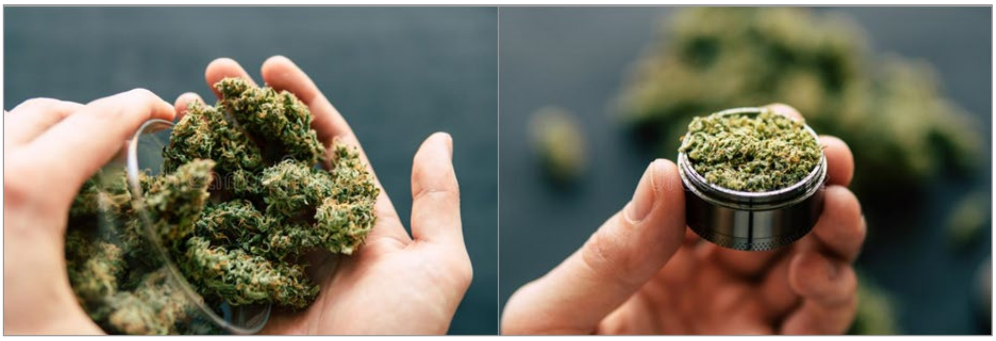

Dry herb 'bud' material | Dry herb 'ground' material

    a. Yes
    b. No

*[If Yes go to Questions 13-21; if No Go to Question 22]*

13. What type of device/s have you used to vaporise dry herb cannabis (flower/plant material)? You may select more than one answer.
    a. A table-top (desktop) vaporiser
    b. A portable (handheld) vaporiser
    c. Other (type response in the textbox below) [TEXT]

14. *[If select table-top option in Q13 include this question]* How often do you vaporise dry herb cannabis in a table-top vaporiser?
    a. Most days
    b. Approximately once a week
    c. Approximately once a month
    d. A few times a year
    e. Once or twice ever

15. *[If select portable option in Q13 include this question]* How often do you vaporise dry herb cannabis in a portable vaporiser?
    a. Most days
    b. Approximately once a week
    c. Approximately once a month
    d. A few times a year
    e. Once or twice ever

16. What brand of vaporiser do you use to vaporise dry herb cannabis? You can provide details on more than one device if you use multiple. [TEXT ANSWER]

17. Approximately how long is the duration of each puff when you inhale vaporised dry herb cannabis (in seconds)? [SLIDE ANSWER – 1 second, 2 second etc up to >10 seconds]

18. At approximately what temperature do you vaporise your cannabis flower/plant material? Please provide your answer as a numerical value and also select the appropriate unit. Temperature can be provided as a range. If unsure leave blank. [TEXT ANSWER]

| | Value | Temperature in degrees Celsius | Temperature in degrees Fahrenheit |
|---|---|---|---|
| Approximate temperature used to vaporise dry herb cannabis | [ ] | ○ | ○ |

19. Approximately how much dry herb cannabis do you vaporise per month? Please provide your answer as an approximate mass, this should only include how much you vaporise and not the amount consumed using other methods (smoking or ingesting). Please also select the appropriate unit. If unsure leave blank.

| | Value | Mass in grams | Mass in ounces |
|---|---|---|---|
| Approximate mass of dry herb cannabis vaped per month | [ ] | ○ | ○ |

20. Once you have finished vaporising your dry herb cannabis do you use the remaining herb material (or AVB) for anything else?
    a. Yes
    b. No

21. *[If Yes to Q20 include this question]* What do you do with the left over dry herb cannabis material after is has been vaporised? [TEXT ANSWER]

**BLOCK 4 - Cannabis Oils/Liquids**

22. Have you ever vaporised cannabis oils or liquids, including cannabis/THC/CBD containing e-liquids in any device including e-cigarettes, vape-pens or dab-rigs? This question is specifically referring to cannabis containing e-liquids, cannabis concentrates which have been dissolved or diluted in a carrier fluid, or cannabis dry herb material which has been extracted directly into a carrier fluid for vaporisation. This excludes the direct use of the dry herb material or undiluted cannabis concentrates.

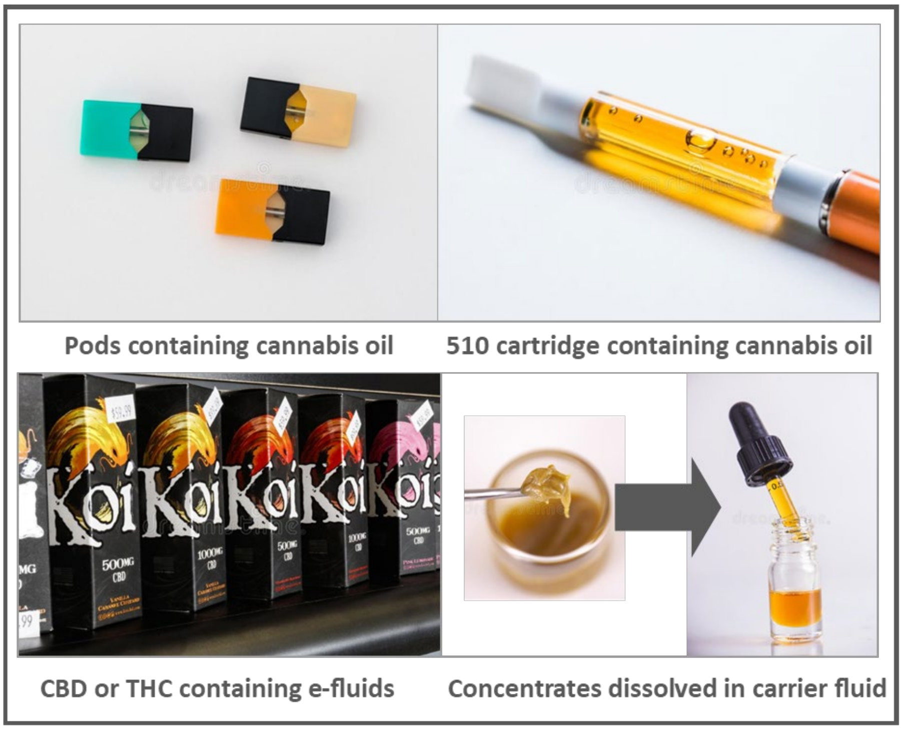

    a. Yes
    b. No

*If Yes go to Questions 23-34; if No Go to Question 35*

23. What type of device/s have you used to vaporise cannabis oils or liquids? You may select more than one answer.
    a. A table-top (desktop) vaporiser
    c. A portable (handheld) vaporiser (e-cigarette or vape pen)
    d. A dab-rig
    e. Other (type response in the textbox below) [TEXT]

24. *[If select table-top option in Q23 include this question]* How often do you vaporise cannabis oils or liquids in a table-top vaporiser?
    a. Most days
    b. Approximately once a week
    c. Approximately once a month
    d. A few times a year
    e. Once or twice ever

25. *[If select portable option in Q23 include this question]* How often do you vaporise cannabis oils or liquids in a portable vaporiser?
    a. Most days
    b. Approximately once a week
    c. Approximately once a month
    d. A few times a year
    e. Once or twice ever

26. *[If select dab-rig option in Q23 include this question]* How often do you vaporise cannabis oils or liquids in a dab-rig?
    a. Most days
    b. Approximately once a week
    c. Approximately once a month
    d. A few times a year
    e. Once or twice ever

27. What brand of vaporiser do you use to vaporise cannabis oils or liquids? You can provide details on more than one device if you use multiple [TEXT ANSWER]

28. Which of the following cannabis oils or liquids have you vaporised? You can select more than one answer.
    a. A purchased cannabis oil (cannabis/terpene flavour)
    b. A cannabis oil made from concentrates (dissolved in terps or thinner)
    c. A cannabis/THC/CBD containing e-liquid (PG or VG based) – unflavoured
    d. A cannabis/THC/CBD containing e-liquid (PG or VG based) – flavoured
    e. Other (type response in the textbox below) [TEXT]

29. Approximately how long is the duration of each puff when you inhale vaporised cannabis oils or liquids (in seconds)? [SLIDE ANSWER – 1 second, 2 second etc up to >10 seconds]

30. At approximately what temperature do you vaporise your cannabis oils or liquids? Please provide your answer as a numerical value and also select the appropriate unit. Temperature can be provided as a range. If unsure leave blank.

| | Value | Temperature in degrees Celsius | Temperature in degrees Fahrenheit |
|---|---|---|---|
| Approximate temperature used to vaporise cannabis oils or liquids | [ ] | ○ | ○ |

31. Approximately how much cannabis oil or liquid do you vaporise per month? Please provide your answer as an approximate volume, this should only include how much you vaporise and not the amount consumed using other methods (smoking or ingesting). Please also select the appropriate unit. If unsure leave blank. [TEXT ANSWER]

| | Value | Volume in millilitres (mls) | Volume in fluid ounces |
|---|---|---|---|
| Approximate volume of cannabis oil or liquid vaped per month | ☐ | ○ | ○ |

32. When you vaporise cannabis containing oils or liquids they are usually in what type of container?

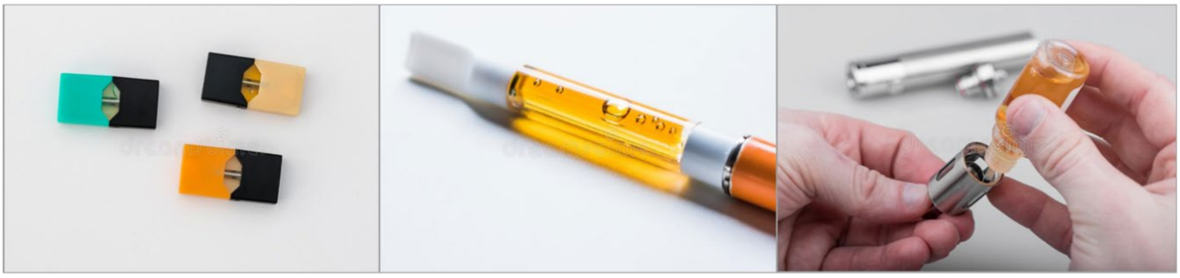

Pods for a pod-style device | A 510 cartridge | Open tank-style device

   a. A pod style container (for example JUUL-style pod which pushes into the vaporiser)
   b. A 510 cartridge (long, thin cartridge which screws into the vaporiser)
   c. An open tank style device which you fill yourself
   d. Other (type response in the textbox below) [TEXT]

33. Where do you obtain your cannabis oils or liquids? You can select more than one option.
   a. From a friend
   b. From a dealer
   c. From a retail store
   d. From the internet (not dark-web)
   e. From the dark-web
   f. I make my own cannabis oils or liquids
   g. Other (type response in the textbox below) [TEXT]

34. *[If select F in Q33 include this question]* If you have made your own cannabis oils/liquids before which methods have you used? You can select more than one option.
   a. Dissolving cannabis concentrates in ‘terps’
   b. Dissolving cannabis concentrates in other e-liquids (PG or VG based)
   c. Dissolving cannabis concentrates in ‘thinners’ or ‘liquidisers’
   d. Extracting dry herb cannabis directly into ‘terps’
   e. Extracting dry herb cannabis directly into other e-liquids (PG or VG based)
   f. Other (type response in the textbox below) [TEXT]

**BLOCK 5 - Cannabis Concentrates**

35. Have you ever vaporised cannabis concentrates (shatter/wax/budder/other) in any device including e-cigarettes, vape-pens or dab-rigs? This question is specifically referring to cannabis concentrates created by extraction of cannabis with solvents and/or heat and excludes vaporisation of the dry herb material or cannabis containing e-liquids or oils.

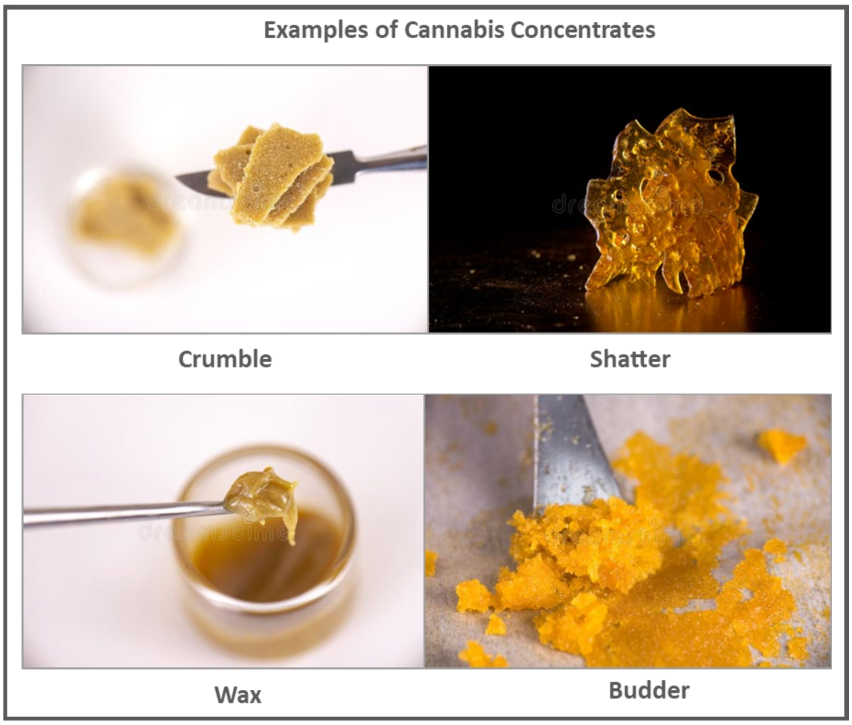

   a. Yes
   b. No

*If Yes go to Questions 36-45; if No Go to Question 46*

36. What type of device/s have you used to vaporise cannabis concentrates (shatter/wax/budder/other)? You may select more than one answer.
    a. A table-top (desktop) vaporiser
    b. A portable (handheld) vaporiser (e-cigarette or vape pen)
    c. A dab-rig
    d. Other (type response in the textbox below) [TEXT]

37. *[If select table-top option in Q36 include this question]* How often do you vaporise cannabis concentrates in a table-top vaporiser?
    a. Most days
    b. Approximately once a week
    c. Approximately once a month
    d. A few times a year
    e. Once or twice ever

38. *[If select portable option in Q36 include this question]* How often do you vaporise cannabis concentrates in a portable vaporiser?
    a. Most days
    b. Approximately once a week
    c. Approximately once a month
    d. A few times a year
    e. Once or twice ever

39. *[If select dab-rig option in Q36 include this question]* How often do you vaporise cannabis concentrates in a dab-rig?
    a. Most days
    b. Approximately once a week
    c. Approximately once a month
    d. A few times a year
    e. Once or twice ever

40. What brand of vaporiser do you use to vaporise cannabis concentrates? You can provide details on more than one device if you use multiple [TEXT ANSWER]

41. Approximately how long is the duration of each puff when you inhale vaporised cannabis concentrates (in seconds)? [SLIDE ANSWER – 1 second, 2 second etc up to >10 seconds]

42. At approximately what temperature do you vaporise your cannabis concentrates? Please provide your answer as a numerical value and also select the appropriate unit. Temperature can be provided as a range. If unsure leave blank. [TEXT ANSWER]

| | Value | Temperature in degrees Celsius | Temperature in degrees Fahrenheit |
|---|---|---|---|
| Approximate temperature used to vaporise cannabis concentrates | ☐ | ○ | ○ |

43. Approximately how much cannabis concentrate do you vaporise per month? Please provide your answer as an approximate mass, this should only include how much you vaporise and not the amount consumed using other methods (smoking or ingesting). Please also select the appropriate unit. If unsure leave blank. [TEXT ANSWER]

| | Value | Mass in grams | Mass in ounces |
|---|---|---|---|
| Approximate mass of cannabis concentrate vaped per month | ☐ | ○ | ○ |

44. Where do you obtain your cannabis concentrates? You can select more than one option.
    a. From a friend
    b. From a dealer
    c. From a retail store
    d. From the internet (not dark-web)

e. From the dark-web
f. I make my own cannabis concentrates
g. Other (type response in the textbox below) [TEXT]

45. *[If select F in Q44 include this question]* If you have made your own concentrates before which methods/solvents have you used? You can select multiple options.
    a. Rosin technique (solventless, hot press)
    b. Solvent extraction with butane
    c. Solvent extraction with ethanol
    d. Solvent extraction with isopropanol
    e. Extraction with carbon dioxide ($CO_2$)
    f. Other (type response in the textbox below) [TEXT]

46. *[If select F in Q44 include this question]* If you have made your own concentrates before which is your preferred/favourite method?
    a. Rosin technique (solventless, hot press)
    b. Solvent extraction with butane
    c. Solvent extraction with ethanol
    d. Solvent extraction with isopropanol
    e. Extraction with carbon dioxide ($CO_2$)
    f. Other (type response in the textbox below) [TEXT]

**BLOCK 6 - Vaping Preferences**

47. If you consume cannabis in multiple ways which of the following is your preferred method?
    a. Smoking
    b. Ingesting
    c. Vaporising
    d. I only vaporise cannabis

48. If you vaporise cannabis in multiple ways which of the following is your preferred method? Please select BOTH your preferred type of cannabis AND your preferred device type or select 'no preferred method'.

| | Type of Cannabis | | | Device Type | | | No preferred method |
|---|---|---|---|---|---|---|---|
| | Dry Herb | Cannabis Oil or Liquid | Cannabis Concentrate | Table-top Vaporiser | Portable (hand-held) Vaporiser | Dab-Rig | |
| Preferred method of cannabis vaporisation | ○ | ○ | ○ | ○ | ○ | ○ | ○ |

49. Why have you decided to vape cannabis products (select all that apply)?
    a. In my opinion it is a healthier alternative to smoking cannabis
    b. It was convenient as I was already a vaper of non-cannabis products
    c. For a better taste experience (more flavour choices)
    d. As it creates a better high for me

e. To replace smoking cannabis in a parallel effort to quit cigarette smoking
f. Because I am a non-smoker
g. This was suggested to me for use of medicinal cannabis
h. Because it is more discreet (stealth vaping)
i. Other (type response in the textbox below) [TEXT]

50. Has vaping cannabis replaced for you one (or more) other mode of using cannabis (smoking, ingesting)?
    a. Yes, it has replaced all other methods (smoking, ingestion)
    b. Yes, it has replace one other method (smoking or ingestion)
    c. No, vaping has not replaced any other method

51. Have you ever heard of synthetic cannabinoids (sometime referred to a legal highs)?
    a. Yes
    b. No

52. *[If YES to Q51 get this question]* Have you ever used synthetic cannabinoids? You may select more than one.
    a. Yes, by vaping in any device
    b. Yes, by ingestion
    c. Yes, by smoking
    d. No

### Supplementary Material 2 – DATA ANALYSIS

**Table SM1:** Chi square contributors for administration method vs reason for use.

| chisq contributors | | | | |
|---|---|---|---|---|
| | | Medicinal & Recreational | Recreational | Medicinal |
| Only Vaporisation | count | 26 | 14 | 6 |
| | expected | 24.29213483 | 15.60898876 | 6.098876404 |
| | contributors | 0.035935111 | 0.079637698 | 0.169413233 |
| Smoking | count | 41 | 55 | 15 |
| | expected | 58.61797753 | 37.66516854 | 14.71685393 |
| | contributors | 5.295186652 | 7.978097362 | 0.005447611 |
| Vaporising | count | 158 | 72 | 32 |
| | expected | 138.3595506 | 88.90337079 | 34.73707865 |
| | contributors | 2.788005978 | 3.213870761 | 0.215665791 |
| Ingesting | count | 10 | 10 | 6 |
| | expected | 13.73033708 | 8.82247191 | 3.447191011 |
| | contributors | 1.013479468 | 0.157163708 | 1.890476539 |
| | | | p value | 0.000850116 |
| | | | df | 6 |
| | | | X^2 | 22.84237991 |

**Table SM2:** Chi square contributors for device type vs cannabis type.

| chisq contributors | | | | |
|---|---|---|---|---|
| | | portable | tabletop | dab-rig |
| dry herb | count | 243 | 55 | |
| | expected | 234.7150538 | 47.26344086 | |
| | contributors | 0.00397492 | 0.015624278 | |
| oil or liquid | count | 32 | 0 | 1 |
| | expected | 25.99193548 | 5.233870968 | 1.774193548 |
| | contributors | 1.388770731 | 5.233870968 | 0.337829912 |
| concentrate | count | 18 | 4 | 19 |
| | expected | 32.29301075 | 6.502688172 | 2.204301075 |
| | contributors | 6.326141528 | 0.963209048 | 127.9750328 |
| | | | p value | 1.59286E-28 |
| | | | df | 4 |
| | | | X^2 | 142.2444542 |

**Table SM3:** ANOVA analysis for mass (g) or volume (mL) of cannabis consumed per month by reason for use.

**Anova: Single Factor**
**Mass Dry Herb Cannabis per Month by Reason of Use**

SUMMARY

| Groups | Count | Sum | Average | Variance |
|---|---|---|---|---|
| Dry Herb Medicinal | 56 | 918.4 | 16.4 | 251.0900909 |
| Dry Herb Med & Rec | 199 | 3391.25 | 17.04146 | 342.3943206 |
| Dry Herb Recreational | 111 | 1188.8 | 10.70991 | 168.9961282 |

ANOVA

| Source of Variation | SS | df | MS | F | P-value | F crit |
|---|---|---|---|---|---|---|
| Between Groups | 2981.847835 | 2 | 1490.924 | 5.401596083 | 0.004879 | 3.020592 |
| Within Groups | 100193.6046 | 363 | 276.0154 | | | |
| Total | 103175.4524 | 365 | | | | |

Post-Hoc Test

| Groups | P-value (t-test) | Significant? |
|---|---|---|
| Med vs Med & Red | 0.813534857 | NO |
| Med & Rec vs Rec | 0.001562144 | YES |
| Med vs Rec | 0.014243213 | YES |

ALPHA

| Test | Alpha |
|---|---|
| ANOVA | 0.05 |
| Post-hoc test | 0.016667 |

**Anova: Single Factor**
**Volume of Liquid/Oil Cannabis per Month by Reason of Use**

SUMMARY

| Groups | Count | Sum | Average | Variance |
|---|---|---|---|---|
| Liquid Medicinal | 11 | 58.5 | 5.318182 | 20.21363636 |
| Liquid Medicinal and Recreational | 70 | 954.52 | 13.636 | 504.9099171 |
| Liquid Recreational | 30 | 427.121 | 14.23737 | 455.8342039 |

ANOVA

| Source of Variation | SS | df | MS | F | P-value | F crit |
|---|---|---|---|---|---|---|
| Between Groups | 723.2870058 | 2 | 361.6435 | 0.809312209 | 0.447845 | 3.080387 |
| Within Groups | 48260.11256 | 108 | 446.8529 | | | |
| Total | 48983.39956 | 110 | | | | |

**Anova: Single Factor**
**Mass of Cannabis Concentrate per Month by Reason of Use**

SUMMARY

| Groups | Count | Sum | Average | Variance |
|---|---|---|---|---|
| Concentrate Medicinal | 32 | 147.85 | 4.620313 | 49.21642893 |
| Concentrate Medicinal and Recreational | 34 | 178.4 | 5.247059 | 65.59787594 |
| Concentrate Recreational | 17 | 59.261 | 3.485941 | 47.79139006 |

ANOVA

| Source of Variation | SS | df | MS | F | P-value | F crit |
|---|---|---|---|---|---|---|
| Between Groups | 35.18173536 | 2 | 17.59087 | 0.315878198 | 0.730053 | 3.110766 |
| Within Groups | 4455.101444 | 80 | 55.68877 | | | |
| Total | 4490.283179 | 82 | | | | |

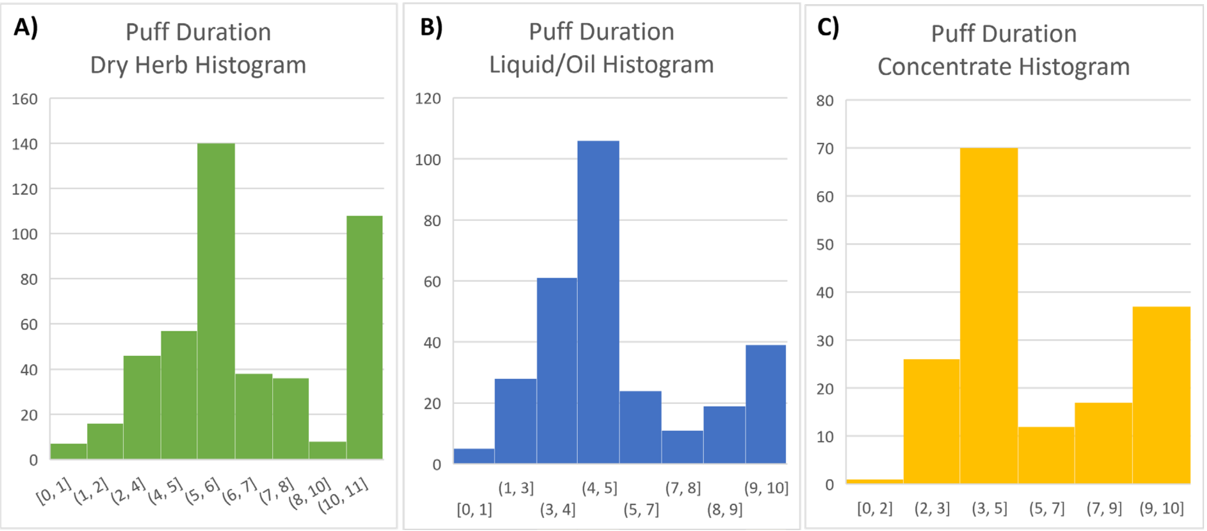

**Figure SM1:** Puff duration histograms (seconds) for **A)** dry herb cannabis (N=456), **B)** cannabis liquid/oil (N=293) and **C)** cannabis concentrate (N=163). The maximum answer available for selection was ten seconds.

**Table SM4:** ANOVA analysis for puff duration (seconds) by type of cannabis.

| | | | | | | |
|---|---|---|---|---|---|---|
| **Anova: Single Factor** | | | | | | |
| **Puff Duration of Different Types of Cannabis - unadjusted data** | | | | | | |
| SUMMARY | | | | | | |
| *Groups* | *Count* | *Sum* | *Average* | *Variance* | | |
| Dry Herb | 456 | 2850 | 6.25 | 6.917582418 | | |
| E-Liquid | 293 | 1501 | 5.122866894 | 6.320468465 | | |
| Concentrate | 163 | 963 | 5.90797546 | 6.701355752 | | |
| ANOVA | | | | | | |
| *Source of Variation* | *SS* | *df* | *MS* | *F* | *P-value* | *F crit* |
| Between Groups | 227.9307693 | 2 | 113.9653846 | 17.0422287 | 5.42E-08 | 3.005627 |
| Within Groups | 6078.696424 | 909 | 6.68723479 | | | |
| Total | 6306.627193 | 911 | | | | |
| Post-Hoc Test | | | | ALPHA | | |
| Groups | P-value (t-test) | Significant? | | Test | Alpha | |
| Dry herb vs e-liquid | 8.59683E-09 | Yes | | ANOVA | 0.05 | |
| E-liquid vs concentrate | 0.001671115 | Yes | | Post-hoc test ( | 0.016667 | |
| Dry herb vs concentrate | 0.152973457 | No | | | | |

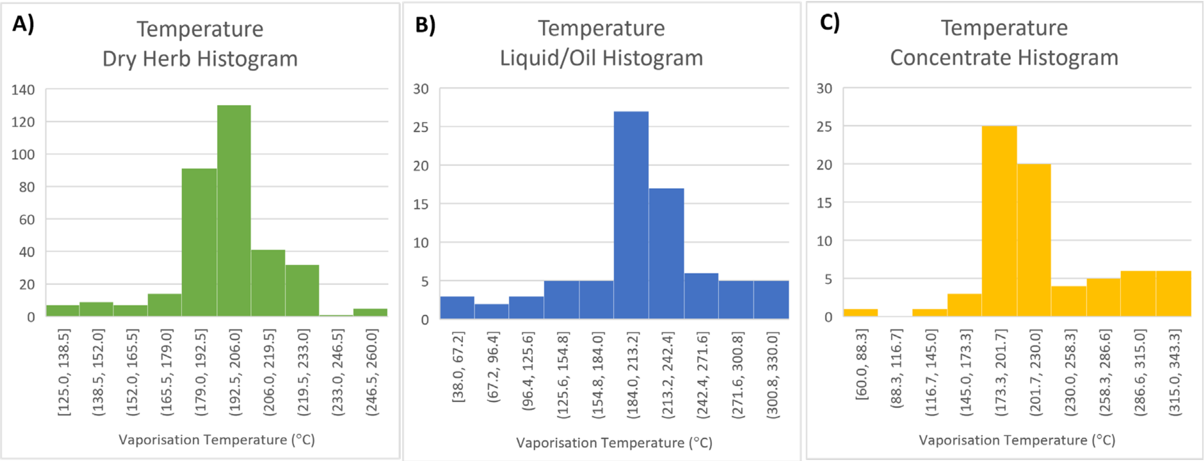

**Figure SM2:** Temperature histograms (°C), outliers removed, for **A)** dry herb cannabis (N=337), **B)** cannabis liquid/oil (N=78) and **C)** cannabis concentrate (N=71).

**Table SM5:** ANOVA analysis for temperature (°C), outliers removed, by type of cannabis.

| | | | | | | |
|---|---|---|---|---|---|---|
| **Anova: Single Factor** | | | | | | |
| **Temperature of Different Types of Cannabis - adjusted data** | | | | | | |
| SUMMARY | | | | | | |
| *Groups* | *Count* | *Sum* | *Average* | *Variance* | | |
| 188 | 336 | 65628.1 | 195.3217262 | 401.2788997 | | |
| | 78 | 16043.3 | 205.6833333 | 3486.203485 | | |
| | 71 | 15662.2 | 220.5943662 | 2512.994825 | | |
| ANOVA | | | | | | |
| *Source of Variation* | *SS* | *df* | *MS* | *F* | *P-value* | *F crit* |
| Between Groups | 39756.82075 | 2 | 19878.41037 | 16.55458787 | 1.11329E-07 | 3.014428773 |
| Within Groups | 578775.7375 | 482 | 1200.779538 | | | |
| Total | 618532.5582 | 484 | | | | |
| Post-Hoc Test | | | | ALPHA | | |
| Groups | P-value (t-test) | Significant? | | Test | Alpha | |
| Dry herb vs e-liquid | 0.008463408 | Yes | | ANOVA | 0.05 | |
| E-liquid vs concentrate | 0.100375342 | No | | Post-hoc test (Bonferroni Correction) | 0.016666667 | |
| Dry herb vs concentrate | 1.02767E-11 | Yes | | | | |

**Table SM6:** ANOVA analysis for temperature ($^{o}C$), outliers removed, of dry herb cannabis and cannabis concentrates by device type (portable, tabletop or dab-rig).

**Anova: Single Factor**

**Temperature of dry herb cannabis vaporisation by device type**

SUMMARY

| *Groups* | *Count* | *Sum* | *Average* | *Variance* |
|---|---|---|---|---|
| Portable Only | 5 | 1055 | 211 | 930 |
| Tabletop Only | 192 | 37394.6 | 194.7635 | 394.0312 |

ANOVA

| *Source of Variation* | *SS* | *df* | *MS* | *F* | *P-value* | *F crit* |
|---|---|---|---|---|---|---|
| Between Groups | 1284.658 | 1 | 1284.658 | 3.171796 | 0.076477 | 3.889589 |
| Within Groups | 78979.96 | 195 | 405.0255 | | | |
| Total | 80264.62 | 196 | | | | |

**Anova: Single Factor**

**Temperature of cannabis concentrate vaporisation by device type**

SUMMARY

| *Groups* | *Count* | *Sum* | *Average* | *Variance* |
|---|---|---|---|---|
| Portable Only | 21 | 4323.3 | 205.8714 | 975.4301 |
| Tabletop Only | 3 | 575 | 191.6667 | 108.3333 |
| Dab-rig Only | 21 | 4701.7 | 223.8905 | 4081.733 |

ANOVA

| *Source of Variation* | *SS* | *df* | *MS* | *F* | *P-value* | *F crit* |
|---|---|---|---|---|---|---|
| Between Groups | 4918.132 | 2 | 2459.066 | 1.018951 | 0.369723 | 3.219942 |
| Within Groups | 101359.9 | 42 | 2413.332 | | | |
| Total | 106278.1 | 44 | | | | |
